# “Running on goodwill and fairydust” - midwives’ experiences of facilitating and delivering local breastfeeding support via Facebook groups: a qualitative descriptive study

**DOI:** 10.1101/2022.10.18.22281224

**Authors:** H. Morse, A. Brown

## Abstract

**Background:** The use of groups for providing and accessing breastfeeding support on Facebook (BSF) is now widespread, including groups aimed specifically at mothers in specific areas. Some of these groups are created and/or facilitated by midwives, but little is known about their motivations or experiences.

**Objective:** To examine how midwives involved in providing breastfeeding support via a local BSF group perceive the value of this provision and what are their experiences of creating these services and of engaging with mothers online.

**Methods:** Semi-structured interviews were conducted with midwives between July-September 2021, and thematically analysed.

**Results:** Three themes were identified in relation to the research question: (1) Imperatives and Value, (2) Goodwill & lack of resources, (3) Community of Practice. Theme 1 described that groups were seen as a necessary part of developing services to meet the needs and expectations of the current generation of mothers, and to improve dire breastfeeding rates. Theme 2 identified that BSF groups are reliant on both midwives and peer supporters working voluntarily, issues of sustainability and frustration at the lack of value placed on their time, skill and investment. Theme 3 highlighted the function of BSF groups as communities of practice, providing opportunities for social learning for all members and personal and professional benefits for midwives.

**Conclusions:** Findings have identified that midwives become involved in delivering online support motivated by a belief in the value of this provision, but feel unsupported by wider services in doing so. Midwives have concerns about the sustainability of this provision regardless of its value whilst it remains reliant on volunteers, and call for further support. Significant benefits for midwives derived from involvement were highlighted which should inform further research, guidance and investment.

## Introduction

Low breastfeeding rates continue to pose a large-scale public health risk [1], and it is well established that access to quality and ongoing support is needed to establish and continue breastfeeding [2]. Online communities in the form of breastfeeding support Facebook (BSF) groups are increasingly used by mothers seeking this support, and research suggests these are highly valued and effective in increasing breastfeeding duration and offering support to meet goals [3]. However, mothers report concerns about reliability [4] and would like further midwifery input into online breastfeeding support to improve reliability and connection with local services [5]. During the COVID-19 pandemic many services turned quickly to online communication [6], but contradictions between national strategy and employer social media policies remain [7]. There are considerable barriers to this service: a lack of training and resources, and perceptions of online engagement with service users as a personal and professional risk [8, 9]. However, evidence suggests that some mothers are currently receiving and highly valuing midwives’ support via Facebook, and that midwives involved in this provision were passionate about doing so, despite largely being unpaid [8].

Whilst such findings present insight into the barriers and facilitators for midwives engaging with online support, no evidence has been identified that explores existing Facebook support provision. Research is needed to understand which challenges have been faced and overcome to facilitate and deliver this provision, how and by whom. Establishing the logistics for midwives of creating, or being involved in moderating them, is fundamental to understanding what is successful, and to meet mothers’ expectations in an increasingly digitally driven social context [9]. Mothers express that collaboration between health professional and peer support is important to them in connecting experiential knowledge, lived experience and trained expertise [10, 5, 8]. Little is known about whether collaboration between any maternity services and third sector breastfeeding organisations is established online, or any logistics of doing so. Sharing solutions to overcoming barriers and identifying ongoing issues will support the development of best practice guidelines to improve outcomes.

Fear of engagement and adverse consequences is a major factor in many midwives’ perceptions of social media use [8, 11], yet midwives involved in delivering online support have fewer concerns and report positive personal and professional benefits [8, 12]. In depth qualitative investigation is needed to understand the context of these differences, establishing to whom the role may be suited and why, or if fear can be mediated by policy, support and training.

Limited evidence suggests a lack of relevant training for midwives in relation to digital skills, social media and digital professionalism, impacting on fear and confidence [8, 11, 13]. Exploring individual experiences of midwives involved in delivering online support is needed to understand perceptions and experience of training and support, to establish what is needed and how to effectively develop and target it. Existing Facebook provision is largely reliant on volunteers [4], even when these are qualified professionals [5], indicative of the wider issues of the under resourcing of breastfeeding support and under valuing of the skills and time of those delivering it. Research to explore insights from those midwives involved will enable recommendations to be made.

Significantly, mothers and midwives engaging in Facebook support report a range of benefits consistent with the conceptualisation of BSF groups as an online community of practice [14], including social connectedness and knowledge acquisition. Further research is needed to understand midwives’ experiences of personal and professional development as a result of online engagement, to inform recommendations for practice.

In order to fulfil these knowledge gaps, we aimed in this investigation to answer the following research question: How do midwives involved in providing breastfeeding support via a local BSF group perceive the value of this provision and what are their experiences of creating these services and of engaging with mothers online?

## Methods

Nine semi-structured interviews were used to collect data on the experiences of midwives involved in creating, facilitating and/or delivering breastfeeding support via a local BSF group. We recruited Registered midwives in the UK (by default over 18, English speaking and able to consent). Convenience sampling was used congruent with the qualitative descriptive design and inclusion criteria, recruiting midwives via Facebook groups aimed at midwives (identified via a Facebook search), with permission sought from group administrators for posting study information to the group or page. The advertisement and link were shared to these groups and shared via Twitter. Ethical approval was received from the University Ethics Committee prior to commencing the study.

Semi structured interviews were considered the most appropriate data collection tool to meet the objectives of this study, aiming to add significant depth to previous findings [8]. Interviews are an ideal way of facilitating a ‘professional conversation’ [15], capturing participants’ perceptions and experiences, and preserving their own language and concepts [16]. The semi-structured format was chosen to ensure participants were offered opportunities to offer insights into the same range of issues, whilst allowing scope for unanticipated issues to be raised [16]. This was key to meeting the study objectives: identifying similarities and differences in perceptions and experiences.

An interview schedule was devised to target the factors involved in setting up and facilitating the BSF group, its perceived value and any personal and professional impacts. Further questions explored the format and facilitation of the group. The interview schedule was semi-structured to allow responsive exploration of the participant’s account, including unanticipated issues, and question wording and order adapted accordingly [16]. See Supplementary material.

All interviews were conducted virtually and recorded with permission, using the Zoom/Teams integrated recorder. Each participant received a £15 Amazon voucher via email in return for their time. All interviews were transcribed and checked against the recording and Identifying information removed from each. Participants were identified by number, ordered by interview date e.g. ‘Midwife 1’.

Thematic analysis using the Braun and Clarke [16] six phase approach was conducted to explore patterns and connections. Firstly, each video interview was re-watched and script read twice for familiarisation with the data. After familiarisation with the data, initial codes (Phase 1) were produced using NVivo v12 (Phase 2), identifying themes (Phase 3).

Experience themes were reviewed in relation to the coded extracts (Phase 4), defined and named (Phase 5). Illustrative extract examples were selected to report results within the final analysis. This was related back to the research questions and relevant literature in the discussion (Phase 6).

## Results

Using thematic analysis three themes were identified in relation to the research question: (1) Imperatives and Value, (2) Goodwill & lack of resources, (3) Community of Practice.

### Theme 1: Imperatives and value

A key issue raised by midwives was the value of providing BSF groups as part of a maternity service, particularly in the context of the COVID-19 pandemic. Midwives in the study reported mixed experiences – some had initiated BSF groups as a necessity, finding support to do so easier to access as a result of the pandemic, whilst others with existing groups felt their impact and value was recognised retrospectively in light of lockdown.

> ‘…there was a face to face service that wasn’t very well set up and didn’t work very well. And when COVID happened … we realised that the women really wanted some sort of parent education and were feeling really vulnerable. And so I said to my boss, let’s just start up a private Facebook page… while we’ve not got anything else on the table and so that’s what we did.’ (Midwife 3)

Midwives whose BSF groups were running prior to the pandemic described them as arising from longstanding gaps in provision, put in place to meet the needs of breastfeeding mothers where health professionals are unable to do so.

> “What’s missing from our services is any care for the woman as a mother… [The BSF group]… provides what missing doesn’t it?” (Midwife 5)

Midwives expressed a passion for supporting breastfeeding mothers as motivation to ‘fill the gap’ and frustration at the limited resources allocated to them in order to do so. A lack of service provision for breastfeeding mothers frequently provided the catalyst for creating and committing to running a BSF group. BSF provision was seen as a way of meeting strategic goals, identifying mothers’ feedback as central to understanding its potential long-term impacts on both experiences and breastfeeding outcomes. Midwives were aware of how mothers’ valued BSF groups and had positive perceptions of the impacts.

> “When we’ve had the [Breastfeeding Friendly Initiative (BFI)] assessments…a theme running through them has been how women talk about the Facebook groups. You know this isn’t just a nice little fluffy bit of extra icing. It’s an absolutely integral part of the care platform. It’s there alongside…the intervention of the midwife [and] is just as important…many of them say I wouldn’t still be breastfeeding if it wasn’t for the Facebook group.” (Midwife 5)

Other benefits included service integration, linking midwifery care and contact with mothers in the community setting enabling increased contact, signposting and information, including between midwives.

> “[A mother] will be asking for a specific help [on the BSF group] and I get a private message for me [from another midwife] to say no, it’s OK, I’ve spoken to her today [in person] and so it was almost like a bit of a hub for us as well…” (Midwife 1)

Midwives also felt that the format encouraged self-efficacy, providing an information resource and source of support trusted as safe and evidence based by women and midwives so some needs could be met without requiring individual input from a professional. They felt this reduced pressure on midwives and providing more cost-effective long-term support enabling women to support and educate one another.

> “[The BSF group] takes some questions off, you know, that women might otherwise be bringing their midwife or their health visitor with. They can post in the group and have some reassurance from other mums or somebody, but I think that the fact that it’s quite heavily moderated by [midwives] means that the information in it is good.” (Midwife 8)

This was also seen as a mechanism to extend the capacity and reach of breastfeeding support services, tapping into the community support and providing evidence based, professionally mediated information that is accessible to more mothers. However, there was a perception again that this was undervalued.

> “Before [the BSF group] our reach for care for specialist information for the women that we looked after, was six a week in our face to face groups… Our Facebook group has around, 1650 women, far bigger than six mums a week, and yet they were paid for their face to face, work, and yet, we’re looking after, a couple of thousand women [without extra resources].” (Midwife 7)

Integrating technology into services was described as imperative to meeting mothers’ needs and expectations, improving their connections with and visibility of midwives. Engaging with social media information seeking was described as an imperative, ensuring accurate information is available, and health professionals are able to address misinformation.

> “The first thing people do is get their phone out for support, isn’t it? They either Google it or they put it on a Facebook page to say, ‘Help, what do I do?’ And if we can get the correct information to people on social media, it’s a lot better than them getting incorrect information in my opinion.” (Midwife 6)

Midwives described how mothers valued access to them via the BSF group, filling the gaps in professional face to face support created by under resourcing. Specifically, input from midwives was highlighted as adding value to the peer support available online, enhancing reliability through evidence-based information.

> “[Mothers] did actually feed that back, that they’d like more availability of the professionals on the [BSF group], so although there is value in the peer support, because it’s that lived experience…they also wanted that authoritative voice in there as well, either confirming or, you know, adding more…so they definitely wanted that [midwife] voice there as well.” (Midwife 1)

Whilst midwives recognised the benefits to the service they were providing, many described the challenges faced by lack of support and often reluctance from management to do so, including a perceived lack of value placed on breastfeeding support and the BSF group. They felt that whilst the pandemic had prompted services to overcome reservations about interacting with service users using social media, enabling new provision and innovative solutions to emerge, guidance was lacking.

> “We’ve all followed our noses doing this, haven’t we, and developed something that we hope works…the main thing is that it’s safe for the women and they trust the service.” (Midwife 5)

In summary, locally linked BSF groups were seen as a necessary part of developing services to meet the needs and expectations of the current generation of mothers, and to improve dire breastfeeding rates. Midwives involved in the provision believe it is incumbent upon maternity and public health services to innovate, to embrace digital communication and to adequately resource and issue guidance for this provision to ensure access is safe and equitable.

### 2. Goodwill and lack of resources

Midwives described a range of BSF group formats, all relying either all or in the most part on them volunteering their own time to facilitate and deliver support. This was embodied by the concept of groups being run on goodwill and ‘fairydust’ (alluding to the lack of renumeration and resources). One midwife described the emotional connection that motivated her continued commitment.

> “It was gonna be finite, that was always the plan. And now it’s been forever… what happens, rightly or wrongly, is that you become slightly obsessed with it, it becomes your little world. I’m sitting there in the evening talking to mums on my group and do that at the weekend and do that when I’m on annual leave. It’s run on goodwill and fairy dust.” (Midwife 3)

The lack of investment and support for BSF group administration was described as having ongoing consequences in terms of future planning, related to the reliance on individuals to maintain online support services in the own time.

> “The NHS is so restrictive I think in terms of introducing new concepts so it had to just be me that took it on the chin if [creating the BSF group] didn’t go as we hoped and I even [struggled] to hand it over as I was leaving. Nobody was willing to take responsibility for it so [running a BSF group] comes down, as a lot of things do in the NHS, to goodwill unfortunately.” (Midwife 1)

The motivation to offer support voluntarily was described as a passion, derived from personal experiences of breastfeeding and breastfeeding support. Midwives were willing to offer their skills voluntarily online to fill the gaps in what they could offer in a paid capacity, borne of a desire to see improvement and prevent harm. They also described the importance of peer support within the BSF group, hinging on time being volunteered by other mothers, either as a parent or trained peer supporter. Trained peer supporters played either a lead or supporting role depending on the format of the BSF group. All agreed collaboration was key to sustainable and quality support.

> “[Online support] has to be a collaborative, you know, thing. It can’t just be us, we can’t just do it because we need the volunteers to run it, but we can’t leave them to do it by themselves. They need back up.” (Midwife 5)

Others felt strongly that the reliance of services on peer supporter to volunteer to run online support provision simply shifted the burden of a lack of resources onto other women, rather than tackling the gaps in funding of professional support.

> “[A peer supporter led format] is probably a way of keeping [the BSF group] going maybe, and taking the load off of [midwives] who are shouldering the load, however, does it diminish the status of the information that we give? And maybe we should step away from that that idea that it’s OK to freeload on women who are at home with their babies? All that really does is passes pressure to another female who is working for free.” (Midwife 7)

Midwives recognised sacrifices made to provide online support and a need for self-protection. The potential benefits of providing online support were limited by the lack of support provided to those facilitating it. The undervaluing of the provision, renumeration for providing it and a lack of recognition of the skill and commitment involved were highlighted. Midwives acknowledged the complex nature of quantifying and qualifying the time spent engaging with mothers online.

> “I run it in my own time. You can meet reluctance [on pay] - like going to a face to face breastfeeding group 100% would be part of hours within work, and again if they were doing a virtual online group we could put it in the diary, but I think stuff like commenting or providing support is one of those things that we just wouldn’t get paid to do.” (Midwife 6)

Midwives also described how they valued the group personally and professionally, underpinning their willingness to give their time and resources voluntarily.

> “My job is huge, it’s not easy to organise and split my time and I think this is added another layer, but because I enjoy it….it gives me that connection with women, it’s a layer that I don’t object to, although I know it’s making my life harder, it’s actually making my life better too.” (Midwife 3)

Providing online support gave midwives a sense of control over the information shared, ensuring women had access to evidence-based information and retaining the ability to remove incorrect or undermining posts.

> “It’s nice to be in a group where you kind of got a measure of control. So, if there’s anything up there that’s really undermining, I can just take it out, you know.” (Midwife 8)

However, a lack of written guidance resulted in midwives becoming and/or feeling individually responsible for ensuring the compliance of the group with breastfeeding standards. This was seen as an additional barrier to wider involvement, centralising the responsibility for maintaining the value of the group.

> [“There’s no written guidance], no it’s all on me, because I’m BFI lead. Obviously it has to be WHO code compliant, you know. And so I’m very aware of that. And if it was somebody else running it, I don’t know if they’d have that on their radar at all, so they might post things that were less BFI friendly.” (Midwife 3)

The lack of recognised framework for providing and facilitating BSF groups was seen by midwives as central to both the ad hoc nature of services and reliance on voluntary commitment. Midwives felt BSF group provision was reliant on them remaining in post, with little support from management. This resulted in concern for the future of the online support they were investing their time and energy in.

> “There is no succession planning for my role ‘cause I’ve just been on annual leave for two weeks and carried on with the Facebook stuff, but my job collapsed while I was away. [The group] would just stop if I went or it wouldn’t stop it would peter out it. It wouldn’t be taken away, but it would just not have the momentum that it has.” (Midwife 3)

This was also described as manifesting as anxiety for those providing the service, requiring both short and long term strategies.

> “If I’m going to be away, I will I let the [other midwives on the group] know and ask them to sort of step up a bit and be mindful I’m not monitoring it, but I do worry and I can’t do it 24/7 forever, can I?” (Midwife 8)

This was recognised a part of the wider issue of undervaluing breastfeeding support and those providing it, in contrast to the expectations of employer support in other midwifery roles, with impact on the perceived sustainability of the role.

> “I don’t think my that my [infant feeding] job is sustainable at all at the moment without goodwill. There isn’t any element of succession planning… and [I had] no meaningful handover.” (Midwife 8)

Midwives discussed group formats as having developed dependent on their own level of commitment and having to learn to manage boundaries to ensure sustainability, particularly in relation to personal messages via Facebook (rather than group posts). Some felt compelled, willing or both to provide this support.

> “It’s actually the private messages that I find more time consuming but I find it really hard to draw the boundaries to be honest…I don’t want anybody to stop breastfeeding because they couldn’t get an answer. Or, you know, access to any support on a Saturday night or something.” (Midwife 8)

Another described having created her own strategies for self-management of the commitment arising for accepting personal messages.

> “I don’t mind [private messages] because it’s not over the top and I think, you know, genuinely if I’m busy or something else is happening that takes priority in my life I have learned that I can not see a message until I’ve got time to see it.” (Midwife 7)

In summary, current local BSF groups are reliant on both midwives and peer supporters working voluntarily. Both do so through personal passion for normalising breastfeeding, a desire to address undermining culture and misinformation to support mothers to meet their goals. Despite this passion, midwives are aware of issues of sustainability and are frustrated by the lack of value placed on their time, skill and investment.

### 3. Community of Practice

This theme demonstrates the ways in which midwives perceived the BSF group as a source of social learning, both for women and themselves. Midwives expressed that the primary intention of setting up a BSF group was to facilitate peer (mother to mother) support whilst offering access to professional support when required, promoting shared learning.

> “I hoped from my experience that it would run itself that you know. I saw these groups that had worked, obviously with a midwife administrator, but that more or less you’d have experienced mums helping less experienced mums. And I will be there just moderating. And that was the hope.” (Midwife 1)

Midwives had experienced development within the group learning that is typical of a community of practice, with knowledge and lived experience being shared, and a commitment to this community learning being demonstrated by the members. Mothers became more expert as a result, relieving pressure on midwives.

> “We’ve seen like exponential growth [of the BSF group] and mothers who we saw in that first year are now very much commentators who peer support other women. And they’re very passionate about doing so, so it’s almost become less time consuming, even though there are more women on there because the information they’re giving is right. So, you skim read the comments now and you’re able to see actually, yeah, she said everything I would.” (Midwife 7)

This was considered an additional reason for identifying as a midwife when posting on the group, adding the authoritative voice to underpinning trust, learning and sharing. Providing evidence-based information and signposting to reliable sources was fundamental to ensuring the same standard of information was shared within the group. Midwives also identified with the community supporting their own learning and development, through observing interactions and prompting further inquiry. This was seen as an additional benefit to the provision.

> “It goes both ways…it’s helping you as a professional in terms of your development, you’re learning but it’s helping the women as well. It ticks every box.” (Midwife 1)

It was also identified that the group served as a learning tool in particularly for newer midwives, but that this was self-selecting and dependent on familiarity with Facebook and motivation to engage. One midwife described how observing peer supporters via the BSF group had actively supported her in acquiring breastfeeding knowledge and boosting professional development, making a strong case for collaboration across support providers with the community.

> “[On the BSF group] the peer supporter goes ‘What about trying this?’ and you think oh, I didn’t even think about that, so it’s like a multi-agency involvement, isn’t it? It’s like anything, the more the more brains you have on one thing, kind of the better. And you definitely learn from each other.” (Midwife 6)

The group provided a catalyst for learning for newer midwives both from women and from the evidence-based information provided by their specialist or more experienced midwife colleagues. Midwives also felt this created a resource of reliable information that could be drawn upon in practice and for professional development. A sense of belonging which motivated women to support each other was identified. This also gave midwives a sense of satisfaction and fulfilment, a key motivator for ongoing involvement.

> “I could see this community growing of women…you’d see kind of the same names come up. They were starting to provide advice and becoming confident themselves that are obviously mums that have breastfed several times. And were just giving the most amazing advice, and I wouldn’t need to do anything. I just kind of let it go. Yeah, that’s spot on and just leave it, And that was amazing.” (Midwife 1)

Midwives described how the BSF group enabled services to reach out to more mothers and families than the traditional support provision. It was felt that this had the potential to create ‘real-life’ breastfeeding communities benefiting more women and babies in future as knowledge and social capital are shared. Normalising breastfeeding culture in an area, as a result of mothers’ belonging to the online breastfeeding support community, was a motivator for midwives, and an effect they reported seeing within the BSF group. This was highlighted particularly in relation to breastfeeding continuation past early infancy.

Midwives also described a sense of personal belonging, of increased social capital and more meaningful relationships, as a result of providing BSF group support over a longer period than usual midwifery care. This was described as personally fulfilling, but also as an investment in the breastfeeding journey that had longer term benefits for mothers and babies, and for the group itself.

> “I think that one of the unexpected values of the group is that connection with mums with older babies, because as a midwife once they’re discharged you don’t have that interaction and the relationship that you build up. But the six weeks that you’re probably giving intense breastfeeding support in the group means that when they’re talking about pumping and going back to work they do come back to us now…[which] is a good thing from my perspective.” (Midwife 3)

It was also considered significant that the BSF groups were seen as ‘belonging’ to the local health service, alongside giving women a sense of ownership of and security within the group itself.

> “With such a poor and breastfeeding rates we have no real breastfeeding community or culture…so having the midwives and health visitors on board and signposting to the Facebook groups so it’s got like that secure feeling to it…that its safe and for everybody.” (Midwife 5)

Midwives described a number of significant connections that they perceived as underpinning the impact and benefits of the local BSF group format. Several highlighted occasions where they had delivered support both face to face and online, contributing to a mothers’ successful breastfeeding journey, fed back to them via the group. This reinforced the value of a connected local service and led to a sense of satisfaction in the job.

> “It’s lovely because you actually see the women in the group months later that you’ve supported in those early days, and I think from a career perspective it’s a really good boost to know that actually, the support obviously worked and she’s still accessing support now, so it’s really nice to see.” (Midwife 6)

Having described the depth of personal passion for, and commitment to providing breastfeeding support, midwives often expressed fulfilment as a result of their investment in the group. One midwife explained her emotional connection to the momentum created within the group.

> “I love [belonging to the BSF group]. I love watching how strong and able women are and passionate and how good information is shared and then re-shared and shared again and you just think yeah, there it is. It’s building, it’s moving forward.” (Midwife 7)

Others identified the importance of the connections between midwives, teams and the women they support via the BSF group in promoting trust and good practice within the community and the wider service.

> “[The success of the group] is about that trust in the [midwifery] team and the relationship that they’ve already built with the team…that’s really important.” (Midwife 3)

Connections between midwives and the BSF groups were also seen as key to mothers’ experience of support, with consistency ensured through ongoing training received by midwives and their engagement with the BSF group.

> “You can post really good research on there and…they’re getting the same message continuously, and that’s one of the things that a lot of women say about breastfeeding is that they’re sick and tired of…being told five different ways to breastfeed a baby, whereas…we share the same resources [on the group] so…women get a consistent approach.” (Midwife 6)

Midwives felt that the most significant connection created by the BSF group was the relationships between mothers. They described ‘real-life’ relationships as being facilitated by the group, and these were seen as positive for mothers health and wellbeing, for extending breastfeeding duration and changing attitudes in the wider local community.

The BSF group was likened to a ‘village’, capable of connecting people with each other, with breastfeeding knowledge and support and anchoring this changing culture within a local community. One midwife described this as using the digital tool to fulfil a need created by changing society.

> “It’s a village. It’s the thing that everyone missed out on as the world expanded and women got pushed into work at the same time as raising children, we lost all of that community and somehow on social media [breastfeeding support] became a village. With all of us saying it’s alright, its normal and you will be ok.” (Midwife 7)
>
> In summary, midwives presented key experiences consistent with conceptualisations of local BSF groups as online communities of practice [14]. Midwives were aware of knowledge acquisition and increased social capital impacting all those engaged with the BSF group, including themselves and the wider healthcare workforce. They also reported a belief in the potential for the CoP to impact the physical local community, generating greater breastfeeding knowledge, shared experience and the possibility of cultural change.

## Discussion

The results identify that midwives offering breastfeeding support via local BSF groups do so because they are passionate about the benefits of this type of support, citing positive impacts on mothers’ breastfeeding experiences and durations and therefore public health. They also confirm previous findings [8], that midwives are providing almost all this support as volunteers, even where the BSF group is linked to their employed role and employer.

Exploration identified that although midwives either deliver or facilitate this support voluntarily, they do so to meet a need that exists as a result of depleted breastfeeding support services, in addition to a belief that online support has benefits in itself. It is clear that this has created a surge in BSF groups linked to maternity services, boosted further by restrictions on face-to-face support during the pandemic, but without clear guidance, or an evidence base establishing best practice. As a result, there is no consensus supporting midwife involvement or collaboration with third sector organisations. The findings have important implications for those considering the integration of online breastfeeding support within maternity services.

Previous research has established that increasing workloads and a lack of capacity, both in midwifery and health visiting, impact on the ability of health professionals to offer breastfeeding support within commissioned services [17]. It is anticipated this will worsen in coming years as ongoing cuts to public health budgets continue [18]. As formula feeding continues to be the UK norm, mothers are increasingly reliant on health professionals for informational and emotional support, with those receiving the help they need more likely to continue past early infancy [3, 19]. Findings echo these issues, highlighting that health professionals passionate about breastfeeding support are frustrated by the restrictions to the service they can provide and keenly aware the impact this has on families.

Breastfeeding support was perceived as a fundamental part of maternity services, and of their role, motivating their willingness to provide this support unpaid, and often without guidance. Describing BSF groups, and breastfeeding support services as a whole, as being run on ‘goodwill and fairydust’, midwives encapsulated the issue of breastfeeding; of mothers, babies and those who support them, as being undervalued. This is a common theme in the literature, which describes how breastfeeding support, despite being a public health responsibility, falls largely on the shoulders of volunteers [20, 4]. Midwives cited reasons for becoming involved with a BSF group and providing support voluntarily, including wanting to ensure no mother stopped breastfeeding through lack of access to timely support and to heal their personal experiences. These reasons also underpinned their willingness to overcome fears in relation to the professional and personal risks of engaging online, mediating these through gaining social media experience and a commitment to service improvement.

Midwives were keenly aware of the perceived risks of online engagement, of the potential for blurred boundaries and issues surrounding digital professionalism. Confidence in their knowledge, ability to access evidence-based information and awareness of quality standards were perceived as mitigating these risks. However, many also reported experiences of moderating the contributions or engagement of other professionals online and perceived a need for further training. Research into the skills and attributes suited to the role, including the development of digital professionalism, was considered central to securing the future of professional online support. This theme is evident in the literature, which recognises that health professionals struggle with the concept of digital professionalism, and that a didactic approach to the changes it poses to practice is not effective [21]. Research is needed to support appropriate training, identifying how to safely adjust the traditional professionalism paradigm to account for the needs and expectations of professionals and services users engaging online [22, 23]. It was evident from the findings that midwives were frustrated by the lack of an evidence base available to underpin frameworks and to justify support and investment by management.

Midwives were aware of receiving mixed messages in clinical practice, where social media use is promoted within national strategy but discouraged in local policy, and this is echoed in the literature [7]. As a result, a lack of support within organisations for designing and delivering BSF groups was common, leaving midwives designing group structures and methods of facilitation. One midwife referred to this as having to “just follow our noses”. As a result, anxieties about the sustainability of the group format, as well as the ongoing impact and implications of being solely responsible for maintaining its function were also common. A sense of overcommitment and personal impacts, including being over capacity and feeling obligated, has been noted in previous studies of health professional moderators [24]. However, in contrast, midwives in this study did not describe an emotional burden, but rather a positive emotional reward from their involvement with and impact on mothers. These findings, in line with the wider literature [5, 12, 14], provide evidence that local BSF groups function as online communities of practice (CoP), conferring informational and social benefits to midwives as well as mothers.

Learning and sharing knowledge within the CoP was a significant motivator for midwives’ involvement. There was a focus on the extended reach of an online CoP, and its ability to confer benefits to the wider midwifery workforce, including students and support workers. The mechanism for this process was often perceived to be exposure to a wider range of mothers’ issues and stories, alongside organic learning from responses to posts by peer supporters and specialist lactation midwives. In effect it enabled midwives to educate their colleagues, enhance signposting and support continuing professional development, and offer a platform for organic learning from mothers shared lived experiences. Midwives perceive a range of personal and professional benefits, including deeper and longer-term connections with women, that motivate their continued participation in the BSF group. Mothers recognise the power of the CoP in normalising the joys and challenges of breastfeeding [3, 14, 25] supporting longer continuation; and the findings confirm that midwives recognise and value this impact, motivating their involvement.

For many midwives the willingness to offer time and skill voluntarily was also a result of improved job satisfaction. Research demonstrates midwives are widely experiencing detrimental impacts to their emotional health and wellbeing in clinical practice [26] the potential for online engagement with women and families to provide increased connection and a sense of fulfilment is therefore a key finding. Increased relational continuity was also reported, a factor known to improve pregnancy and birth outcomes and women’s experiences, in addition to improved fulfilment for midwives [27]. Continuity of care for all women is a vision which has been adopted across national maternity strategies [28, 29], yet never achieved as a result of widespread systemic barriers [30]. Some improvements could be made through increased relational continuity through integration of online midwifery moderated support groups [12] as reported in the findings.

Collaboration with others was also considered key to maintaining and sustaining an online service, and formats were all linked formally or informally to trained peer support. Mothers seek access to trained peer support, and this provision is recommended by the WHO [31] and reflected in UK guidance [32]. However, there are no specified models for delivering this support within services, resulting in heterogeneity of services and inequitable access [33]. Midwives cited institutional and managerial barriers to engaging with third sector breastfeeding organisations in their efforts to sustain groups; a need confronting to a hierarchical NHS culture. These are evident in the literature as conflict arises in a hierarchical system where greater value is given to professional contributions [34, 35]. Findings of the study identify that designing and facilitating peer support involvement in maternity service BSF groups centred on this issue; how to facilitate integration and maintain service oversight with a level of safety and acceptability to all.

BSF groups were seen to offer maternity services a format for improving digital literacy and investing in the digital transformation of NHS support, with minimal cost implications. Midwives believed innovation to meet the expectations of digital support current and future mothers have is needed, sharing the vision put forward by national strategy [36]. They expressed ways in which the services they were running had the potential to improve care, cut costs and increase efficiency [37], and yet lacked funding and investment. Reluctance to invest in digital services, breastfeeding support and failure to recognise the personal investment made by midwives in delivering social media support were seen as systemic, embedded in the organisational culture of the NHS. As a result, midwives felt unsupported in developing digital support services.

Policies, protocols and procedures underpin the delivery of care within the NHS. These and the discourse associated with developing and delivering them are fundamental to an organisational culture of safety [38]. To deliver safety to service users and staff, this culture must comprise of openness, leadership, support, communication and the integration of risk management, and continuously seek to minimise harm and optimise care [39]. Midwives have created and facilitate local BSF groups to support women and to improve experiences and outcomes. However, the lack of leadership and policy support raises safety concerns, including questions of training, quality, sustainability and access, which were identified by both mothers and midwives. Volunteer pathways and solutions to reliance on midwives working unpaid in the role to sustain services are urgently needed.

### Limitations

This study does have its limitations. Whilst online, video-based interviews have become an increasingly popular tool, they also have limitations, effecting many aspects of the research process. Obtaining written consent via digital methods, the digital literacy of the participants and researcher and considerations of platform incongruence needed to be considered (Zoom/Teams) [40]. In addition, interpreting the non-verbal communication that underpins analysis can be limited by video technology, particularly as being distracted by the image of oneself on-screen and a lack of eye contact is common [41]. However, there are numerous benefits, reducing travel time and costs and offering a convenient, accessible way for geographically disparate participants to take part [40].

With regard to sampling, only interested and motivated participants may have taken part, omitting the views of those who may have more negative experiences of BSF group facilitation. The sample size was also limited by time and resources. Further research may want to explore the experiences of peer supporters involved in these specific BSF groups, to understand the wider impact of the different approaches and levels of midwifery involvement.

It is recognised that the participants were aware of the researcher’s background as a midwife and interest in social media support, and this may result in confirmation bias [42]. As social media use by midwives is presented as professionally problematic by educators, employers and professional bodies, only those midwife participants happy to be identifiable to another midwife conducting the research would have agreed to take part, despite the anonymising of data post collection. However, open questioning and building a positive rapport with those electing to take part promoted trust in confidentiality, enabled high validity, encouraging detailed answers and exploration.

## Conclusion

This study has demonstrated the impact that a lack of research into and support for the BSF group format has had on individual midwives and equity of the service. Without a clear evidence base, midwives in different areas have developed approaches to delivering breastfeeding support via social media in isolation. Findings have confirmed that without established frameworks and recognition of the significant contribution made to breastfeeding support by BSF groups and the commitment of volunteers, midwives fear these services will become unsustainable despite their impact on mothers and public health. Limitations aside, this research has confirmed the significance of BSF groups in maternity services and established the importance of disseminating this evidence base to underpin investment and development. Further research is needed to develop insights into the barriers and facilitators to developing this service, including group formats and logistics.

## Data Availability

All data produced in the present study are available upon reasonable request to the authors

